# Genome x Environment analysis of Sudden Unexpected Infant Death unveils etiologic heterogeneity and strong cannabis and genetic disease risks

**DOI:** 10.1101/2025.11.26.25341098

**Authors:** Stephen F. Kingsmore, Gretchen Bandoli, Daniel C. Helbling, Rebecca Baer, Eric Blincow, Bryant Cao, Erwin Frise, Alaina Heinen, Laura Jelliffe-Pawlowski, Erica Sanford Kobayashi, Lucita Van Der Kraan, Hugh Kwon, Rishona Lavy, Barry Moore, Danny Oh, Scott Oltman, Eric Ontiveros, Liana Protopsaltis, Mark Yandell, Christina D Chambers

## Abstract

Sudden Unexpected Infant Death (SUID), the third leading cause of infant death, has increasing incidence and multifactorial etiology. Identification of preventative interventions has hitherto been hindered by etiologic studies limited to genetic or environmental effects in isolation. Here we report a multifactorial genome x environment analysis of SUID risk. Births in San Diego County California from 2005–2018 were linked to hospital discharge summaries and death files, yielding 212 SUID cases and 620,392 infants alive at age 1 year. Whole genome sequencing (WGS) identified probable and possible genetic etiologies in 16% and 48% of SUID cases, respectively. Genetic risks were extremely heterogeneous with 144 loci contributing 173 risks in 57% of SUID cases. Genetic risk was very strong (Prevalence Risk Ratio, PRR >99) or strong (PRR 3.7 – 99) in 12% and 34% of SUID cases, respectively. Six of sixteen significant environmental risks lost significance when SUID cases without strong or very strong genetic risk were compared with infants alive at age 1 year, while SUID risk associated with prenatal cannabis increased from adjusted hazard ratio (aHR) 3.7 to 6.0, other substance abuse from aHR 2.6 to 3.5, and black race from aHR 1.9 to 2.5. Thus, genome x environment analysis of a large cohort unveiled etiologic heterogeneity and hidden SUID risks, highlighting cannabis and genetic diseases as strong risk factors. Since preventative or therapeutic interventions were available for 83% of genetic risks, newborn screening by WGS has potential for substantial SUID reduction. Educational campaigns for SUID should emphasize perinatal cannabis avoidance.

**One Sentence Summary:** Multifactorial genome x environment analysis of a large cohort identified heterogeneous etiology in Sudden Unexpected Infant Death and unveiled strong risks from prenatal cannabis and genetic diseases.

## Introduction

Sudden Unexpected Infant Death (SUID) is death at <1 year of age that is sudden and unexpected^1,2^. SUID is the third leading cause of infant death in the United States (US). As defined by the American Academy of Pediatrics (AAP), SUID includes both explained and unexplained sudden infant deaths^2^. As defined by the US Centers for Disease Control and Prevention (CDC), SUID requires that the cause was not obvious before investigation^1^. The 10th revision of the International Classification of Diseases (ICD-10) has three codes that together comprise SUID in CDC national vital statistics reports: R95 encoding Sudden Infant Death Syndrome (SIDS, sudden infant death unexplained by autopsy, examination of the death scene, and review of the clinical history), R99 encoding other ill-defined, unspecified, unknown or undetermined causes of infant mortality, and W75 encoding accidental infant suffocation and strangulation in bed. Knowledge of SUID etiology shapes public health policy and priorities for investments in surveillance, intervention and medical research. For example, association of prone sleeping position with SIDS was identified in the 1970s and led to a supine sleep recommendation by the AAP in 1992 and the Back to Sleep/Safe to Sleep campaign in the United Kingdom and US in 1991 and 1994, respectively^3–5^. Partly as a result, the prevalence of SUID in US infants decreased from 180/100,000 in 1979 to 86/100,000 in 2011^6–8^. However, SUID prevalence has risen since, to 101/100,000 in 2022^9^. During this period, tremendous advances in molecular understanding of disease etiopathogenesis have led to improved outcomes through precision medicine but have not extended to SUID^10^. While WGS or whole exome sequencing have led to molecular insights into the etiopathogenesis in individual SUID^10–15^, synthesis with traditional epidemiologic knowledge of SUID risk has not occurred to create a unified understanding that informs new precision preventative interventions.

Previous studies have shown SUID risk to be both highly multifactorial and heterogeneous between individuals^10–28^. SUID risk factors have been mapped to four domains: maternal and perinatal environmental exposures, social factors, and lifestyle factors (herein together referred to as **E**), and infant monogenic and oligogenic factors (herein referred to as **G,** Fig. 1a). Hitherto, risk factors have been examined individually or by domain, offering an incomplete and potentially confounded picture of SUID etiology. Current SUID risk models are likely to have failed to distinguish risk factors from different domains that are either intermediates in causal chains (and act to modify a direct risk factor), colliders, or confounders^29–31^. As a result, some risk factors may be invalid or underweighted. Synthesis across **E** and **G** SUID risk domains has the potential to inform a unified etiologic understanding with valid inference of causal risk and accurate effect size estimation. Integrative analysis may also allow the definition of homogeneous etiologic subsets with precision (i.e. individualized) strategies for SUID prevention that address persistent disparities in rates^10,17,18,25^. Integrative analyses, however, are challenging both because of etiologic heterogeneity between SUID subgroups and due to a dearth of validated methods for **E** x **G** modeling in an uncommon, highly heterogenous outcome such as SUID.

**Fig. 1:**
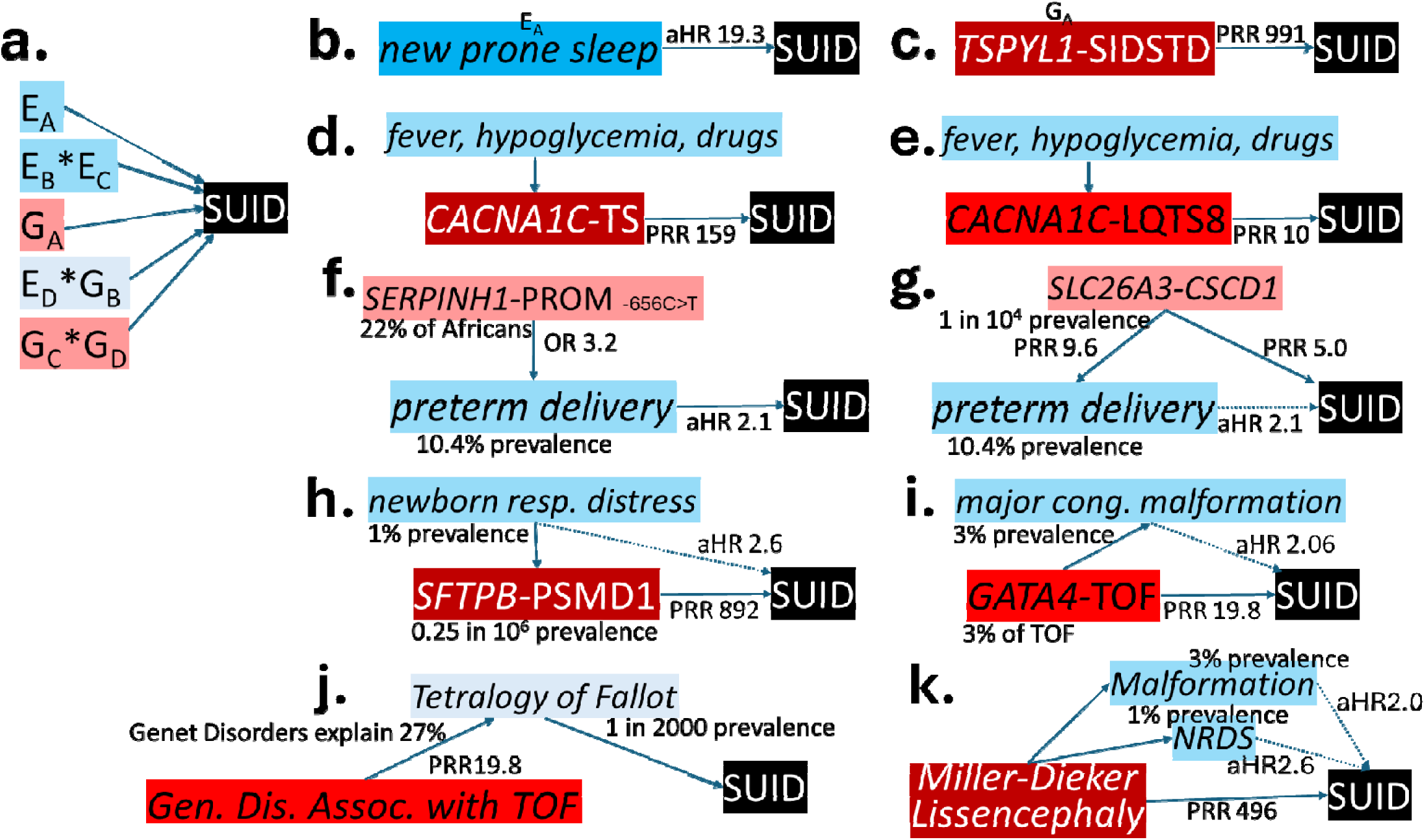
Graphs of integrated G * E models of causal association of SUID. Arrows with solid lines represent direct associations and those with dotted lines represent associations subsumed by a direct association. Birth prevalences are indicated. Risk magnitudes are indicated by prevalence risk ratios (PRR) for G factors and cohort adjusted hazard ratios (aHR) for E factors (from Table 1). **a** Interactions of SUID risk genetic (G, red) and non-genetic factors (E, blue). E_A_ and G_A_ indicate factors with independent, direct causal association with SUID (not confounded by other factors). E_B_, E_C_, E_D_, G_B_, G_C_, and G_D_ represent factors with direct but dependent causal association with SUID (potentially confounded or amplified by other factors). Examples shown are new prone sleep (**b**), *TSPYL1*–SIDS with dysgenesis of the testes (SIDSTD, **c**), *CACNA1C*–Timothy syndrome (TS, **d**), *CACNA1C*–Long QT syndrome 8 (LTQS8, **e**), *SERPINH1*–susceptibility to preterm premature rupture of the membranes (SPROM, **f**), *SLC26A3*–Congenital secretory chloride diarrhea 1 (CSCD1, **g**), *SFTPB*–pulmonary surfactant metabolism dysfunction disorder 1 (PSMD1, **h**), *GATA4*–Tetralogy of Fallot (TOF, **i**), Tetralogy of Fallot of all causes (**j**), and Miller-Dieker Lissencephaly (**k**). These are discussed in Supplementary Results. Fever, hypoglycemia, and various drugs exacerbate cardiac dysrhythmias in TS and LTQS8. Other than nicotine, alcohol, and substance abuse disorder these have not been shown to have an association with SUID. Prematurity can have either a direct association with SUID (as in SPROM) or indirect (as in CSCD1). Newborn respiratory distress syndrome and major congenital malformations can have either direct or indirect associations with SUID (as in PSMD1 and TOF, respectively).

**Table 1:** Adjusted hazard ratios (aHR) and 95% confidence intervals (CI) for maternal, infant and environmental characteristics of SUID cases compared to 620,392 infants alive at age 1 year. Comparison groups were all SUID cases (n=212), SUID cases without very strong genetic risk (n=186), and SUID cases without strong or very strong genetic risk (n=114). Models adjusted for maternal race/ethnicity, age, education, body mass index, prenatal care, payer source, and smoking. Each factor from individual models. aHR in bold were significant (p<.05).

| Factor | Characteristic | All SUID<br>aHR (95% CI) | SUID Without Very<br>Strong G Risk aHR<br>(95% CI) | SUID Without Very<br>Strong or Strong G<br>Risk aHR (95% CI) |
| --- | --- | --- | --- | --- |
| Race/ethnicity | White | reference | Reference | reference |
|  | Hispanic | **0.6 (0.4, 0.8)** | **0.6 (0.4, 0.8)** | **0.6 (0.4, 0.9)** |
|  | Black | **1.9 (1.1, 3.0)** | **1.9 (1.2, 3.2)** | **2.5 (1.2, 4.7)** |
|  | Asian | 0.6 (0.3, 1.1) | 0.6 (0.3, 1.2) | 0.4 (0.2, 1.3) |
|  | Other, multiple or unknown | 0.9 (0.6, 1.4) | 1.0 (0.6, 1.7) | 0.8 (0.4, 1.6) |
| Age | <18 | 1.1 (0.5, 2.5) | 0.9 (0.4, 2.3) | 0.6 (0.1, 2.5) |
|  | 18-34 | Reference | Reference | reference |
|  | 35+ | **0.5 (0.3, 0.8)** | **0.5 (0.3, 0.8)** | **0.4 (0.2, 0.8)** |
| Education | <12th grade | **1.6 (1.2, 2.3)** | **1.6 (1.1, 2.3)** | **1.8 (1.1, 2.8)** |
| Body mass index | Overweight or obese | **1.6 (1.2, 2.2)** | **1.5 (1.1, 2.1)** | 1.6 (1.0, 2.3) |
| Prenatal care | Inadequate | **1.8 (1.3, 2.5)** | **1.9 (1.3, 2.8)** | 1.4 (0.9, 2.6) |
| Payer source | Private | Reference | Reference | Reference |
|  | Public | **1.8 (1.3, 2.5)** | **2 (1.3, 2.8)** | 1.6 (0.9, 2.2) |
|  | Other | 1.4 (0.8, 2.6) | 1.3 (0.7, 2.6) | 1.1 (0.5, 2.6) |
| Parity | Multiparous | **1.5 (1.1, 2.0)** | **1.6 (1.1, 2.2)** | **1.6 (1.1, 2.5)** |
| Mental health | Anxiety | 1.2 (0.6, 2.5) | 1.4 (0.7, 2.9) | 0.9 (0.3, 2.9) |
|  | Depression | **2.1 (1.2, 3.8)** | **2.4 (1.3, 4.4)** | **2.4 (1.1, 5.2)** |
| Substance related<br>diagnosis | Other substance related diagnosis | **2.6 (1.6, 4.6)** | **3.0 (1.7, 5.2)** | **3.5 (1.7, 7.3)** |
|  | Cannabis related diagnosis | **3.7 (2.1, 6.7)** | **4.4 (2.5, 7.9)** | **6.0 (3.0, 12.0)** |
|  | Nicotine | **2.9 (1.7, 4.7)** | **2.8 (1.6, 4.9)** | **2.4 (1.5, 5.1)** |
| *Comorbidities* | Preexisting hypertension | **2.4 (1.2, 4.7)** | **2.4 (1.2, 4.9)** | 1.9 (0.7, 5.3) |
|  | Preexisting diabetes | 0.5 (0.1, 3.5) | 0.6 (0.1, 3.9) | 0.9 (0.1, 6.6) |
|  | Gestational diabetes | 1.1 (0.7, 1.7) | 1.2 (0.7, 1.9) | 1.3 (0.8, 2.4) |
|  | Gestational hypertension | 1.5 (0.8, 3.0) | 1.6 (0.8, 3.2) | 1.9 (0.9, 4.5) |
|  | Preeclampsia | 0.9 (0.4, 1.7) | 0.9 (0.5, 2.0) | 0.8 (0.3, 2.2) |
| *Pregnancy complications* | Preterm delivery | **2.1 (1.4, 3.0)** | **2.0 (1.4, 3.0)** | **2.1 (1.3, 3.4)** |
|  | Very preterm delivery | **3.4 (1.6, 6.8)** | **2.8 (1.3, 6.4)** | 1.5 (0.4, 6.2) |
| *Infant characteristics* | Major structural malformation | 1.5 (0.7, 2.8) | 0.9 (0.3, 2.2) | 1.1 (0.5, 2.5) |
|  | Respiratory distress syndrome | **2.5 (1.4, 4.4)** | 1.8 (0.9, 3.7) | 1.9 (0.8, 4.6) |

We have previously performed separate multifactorial analyses of SUID risk for **E** and **G** domains^16–18^. We evaluated 45 potential SUID risk **E** factors in 211 SUID cases and 482,829 infants alive at age 1 year in San Diego County (SD) California (CA) between 2005-2017^18,23,32^. Nine E factors were significant SUID risks (multiparity, maternal depression, prenatal maternal cannabis and nicotine use, other substance-related diagnoses, preexisting hypertension, preterm delivery, major congenital malformation, and respiratory distress syndrome). While these factors occurred commonly in populations (prevalence >1%), they had relatively weak effects (adjusted hazard ratios, aHR 1.4–2.7). Each has been replicated^4,7,18,20,23–25^.

We also evaluated infant death risk **G** risk factors in 112 infant deaths and 434 infants alive at age 1 year among critically ill infants receiving intensive care in SD between 2015-2020^16,17^. G risk factors were identified by phenotype-informed whole WGS, which detects almost all ∼10,000 known single gene and structural variant diseases^33,34^. Genotypes that were either highly likely to cause disease (pathogenic (P) and likely pathogenic (LP) variants) or possibly likely to cause disease (variants of uncertain significance, VUS, that matched phenotypes in corresponding death certificates and medical records) were identified in 41% of infant deaths, of which 83% of genes had shown prior association with childhood death^17^. Genetic heterogeneity was extreme, with 96% of diseases being unique to a single infant death. In contrast, only 3% of 434 critically infants alive at age 1 year had these genetic diagnoses^17^. These results were consistent with prior genomic sequencing studies in SUID or overlapping infant death outcomes^11–15,21,22,25–28^. In contrast to E risk, most SUID risk **G** factors were very rare in populations (prevalence <1 in 10,000) but conveyed strong risk.

Here we report multifactorial **G** x **E** analysis of SUID risk in 620,604 births and 212 SUID cases in SD from 2005-2018. We identified **G** factors by WGS of SUID cases and examined 45 **E** factors across maternal, infant and environmental domains in the entire cohort.

## Subjects and Methods

### Experimental Design and Research Participants

The study design is outlined in Fig. S1. We identified all SD live births between 2005-2018 using the Study of Mothers and Infants (SOMI), a comprehensive dataset of administrative health records of all CA births^18,32^. SOMI includes >6 million mother-baby dyads with discharge records (maternal: one year prior to birth through one year post-delivery) linked to infant birth certificates and death records for any infant who died in CA. Birth certificates were linked to ICD codes from hospital discharge, emergency department, and ambulatory surgery records by maternal and infant dates of birth, zip code, delivery hospital, pregnancy complications and outcomes, and payer. CalEnviroscreen 3.0 and other publicly available geographically mapped data were linked through zip codes at delivery. All infant deaths in SD between 2005-2018 were identified, and 212 SUID cases with archived CA newborn screen (NBS) dried blood spots (DBS) available were identified using ICD-10 codes R95, W75, and R99 from infant death certificates and records or hospital discharge summaries. The study was approved by the Committee for the Protection of Human Subjects within the CA Health and Human Services Agency and the University of CA-SD Institutional Review Board.

### Whole genome sequencing

WGS was performed on 212 de-identified SUID cases as described^35^. Briefly, genomic DNA was isolated from five 3mm^2^ NBS DBS punches with the DNA Flex Lysis Kit (Illumina). Sequencing libraries were prepared with DNA PCR-free kits (Illumina). Libraries with concentration >3nM were sequenced (2×101 nucleotide) on NovaSeq 6000 instruments (Illumina). WGS quality controls were Q30 ≥80%, error rate ≤3%, and >120Gb sequence/sample. WGS were aligned to GRCh37 and variants identified and genotyped with DRAGEN v3.9 (Illumina). Structural variants were filtered to retain those affecting coding regions of Mendelian Inheritance in Man (MIM) disease genes and with allele frequencies <2% in the RCIGM database. Additional quality controls included: identity tracking by CODIS short tandem repeats by capillary electrophoresis and *in silico* from WGS; <15% duplicates; >98% aligned reads; Ti/Tv ratio 2.0-2.2; Hom/Het variant ratio 0.40-0.61; >90% of MIM genes with >10-fold coverage of all coding nucleotides; sex match; coverage uniformity by GC bias; standard deviation of coverage normalized to average coverage; and the total length of GRCh37 with read coverage. Genetic variants were interpreted according to standard guidelines by clinical molecular geneticists with ACE, GEM, Transformer, and Enterprise software (Fabric Genomics) using the variant call file (vcf), human phenotype ontology (HPO) terms from death certificates and the term Death in Infancy (HP:0001522).

### Calculation of G risk in SUID cases

WGS genotypes were retained if they contained P or LP variants, or VUS with appropriate mode of inheritance (MOI) in MIM genes associated with a disease that had been reported to cause death in fetuses, infants or children, and if their population frequency was consistent with SUID prevalence. Where more than one disorder was associated with a gene, the disorder most likely to cause SUID was utilized. We calculated two SUID-associated genetic disease genotype frequency ***f*** cutoffs and used the larger. We first calculated ***f*** with the model^35^:

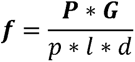

Where *P* was the US prevalence of SUID (101/100,000)^8^, *G* the proportion of SUID attributable to all genetic disorders (set at 0.33), *l* the median SUID genetic locus heterogeneity (set at 100), *p* the median likelihood of SUID for that disorder (0.9, Table S1), and *d* the median genotype heterogeneity of SUID loci (set at 20). With these values, ***f*** = 1.9 × 10^−7^. Secondly, we calculated ***f*** based on aggregated WGS of 807,162 million adults (gnomAD v4.1). Genotypes with high expressivity and penetrance for SUID risk should not be observed in gnomAD^35^. Therefore, we calculated the Wilson score 99.9% confidence interval (CI) upper bound for zero occurrences in 807,162 subjects (8.2 × 10^−6^). SUID cases with genotypes with P or LP variants and ***f*** <8.2 × 10^−6^ were categorized as probable genetic etiology, and those with genotypes with VUS and ***f*** <8.2 × 10^−6^, or with P/LP variants and ***f*** <3.6 × 10^−5^ as possible genetic etiology (implying *p* <0.25).

We evaluated birth prevalence, penetrance, likelihood of infant death, and SUID prevalence risk ratio (PRR) for 107 genetic diseases (Table S1), where: *PRR* = *p/P*. These examples informed conversion of probable or possible genetic etiology categories to quantitative SUID PRRs for each disorder. PRRs were calculated for SUID cases based on two assumptions: Firstly, the PRR for a disorder of possible genetic etiology was assumed to be the square root of the PRR for that disorder when of probable genetic etiology. Secondly, for SUID cases with >1 genetic disorder, we combined PRRs by calculating the product of individual PRRs. We converted PRRs of SUID cases into three categories: Those with very strong genetic risk (combined PRR >99, or 10% SUID risk), where E factors were considered noncontributory to SUID; strong genetic risk (PRR 3.7 – 99), where G x E interactions contributed to SUID outcomes; and weak genetic risk (PRR <3.7), where E factors outweighed G factors in SUID outcomes.

### SUID risk factors in maternal, pregnancy, or infant E domains

SUID-risk maternal and infant characteristics and environmental pollutants were primarily selected from our prior analysis of SUID^18^. Maternal factors included race/ethnicity (non-Hispanic White, Black, Asian, Hispanic, or other/multiple/unknown), nativity (US born or foreign born), age (<18, 18-34, >34), education (<12^th^ grade or 12^th^ grade or higher), pre-pregnancy body mass index (≥25 kg/m^2^), prenatal care(inadequate or adequate+), and payer source for delivery (private, public, other). All queried factors were pre-specified.

### Statistical Analysis

We assessed univariate associations between each E risk factor and SUID vs. alive at one year with chi-square, Fisher’s exact or t-tests, as appropriate. Factors with univariate probability ≤0.1 and count ≥5 were included in multivariable regression. The significance was set to 0.1 due to the small strata for SUID (n=212). Adjusted hazard ratios (aHR) and 95% confidence intervals (CI) were calculated by Cox multivariable regression. Age at death (days) was the unit of time. Non-SUID deaths were censored at the day of death, and infants alive at one year were censored at one year. We first focused on a maternal model, regressing SUID on maternal race/ethnicity, nativity, age, education, pre-pregnancy BMI, prenatal care, and payer source. We performed models with and without imputed variables, and, observing no marked differences, we used imputed variables in subsequent analyses. We then constructed individual models for each maternal, infant and environmental factor with univariate significance, each adjusted for the aforementioned maternal characteristics. We considered that maternal smoking, a well-documented SUID risk factor, may confound variables in the prenatal and infant models. Therefore, we adjusted each model for maternal nicotine use (except the model estimating maternal nicotine risk). Individual models for each significant factor were calculated in comparisons with all SUID cases (n=212), SUID cases without very strong genetic risk (n=186), and SUID cases without strong or very strong genetic risk (n=113, Fig. S1). Sensitivity analyses were comparisons of all SUID cases, SUID cases without probable genetic etiology, and SUID cases probable or possible genetic etiology. For G risk factors, CI of binomial proportions were calculated with Wilson score 95% CI and differences in categorical variables were calculated by Fisher’s exact test. Analyses were conducted in SAS, version 9.4 (SAS Institute, Cary, NC).

## Results

We report findings from comprehensive, quantitative analyses of environmental and genomic (**E** x **G)** SUID risk in 212 SUID cases, >600,000 infants alive at age 1 year (Fig. S1). There were 45 **E** factors representing maternal, infant and environmental domains, and all genetic disorders associated with infant death identified by WGS.

Of 620,606 live births in SD from 2005-2018, 2,686 died in infancy (infant mortality rate 433/100,000, range 355-509/ 100,000)^48,49^. Of these, 227 (8.4%) were SUID (SUID rate 36.6/100,000). Of 227 SUID, 212 had archived newborn screening dried blood spots. Death certificates of the 212 SUID cases listed ICD-10 code R95 (SIDS) in 176, R99 (Other Ill-Defined and Unspecified Causes of Mortality - Unknown/ Undetermined) in 14, and W75 (Accidental Suffocation and Strangulation in Bed) in 22.

### Whole Genome Sequencing (WGS) of SUID cases

SUID G risk was assessed in 212 SUID cases by WGS clinical interpretation using the phenotype Death in Infancy (HP:0001522) and other phenotypes from death certificates. Following interpretation, 188 rare genotypes were retained in 121 (57%) of the 212 SUID cases (Table S2). They mapped to 144 single locus genetic disorders with known associations with infant death.

### Identification of probable genetic etiologies in individual SUID cases

The 188 genotypes were categorized to reflect either probable or possible genetic etiology. Thirty-four (16%) SUID cases had probable genetic etiology based on 36 rare genotypes (*f* <8.2 × 10^−6^) in 33 loci with MOI for genetic diseases that can cause infant death. In 30 of the 34 cases, genotypes met criteria for molecular diagnoses (P and LP variants). The remaining four cases had rare VUS associated with genetic diseases whose presentations overlapped sufficiently with death certificate phenotypes to upgrade them: One had death certificate phenotypes of SUID, coarctation of aorta, and patent ductus arteriosus. Aortic coarctation occurs with prevalence of 4/10,000 births^50^. The rare VUS was *SMAD6* c.778T>C (p.Cys260Arg). *SMAD6* is associated with autosomal dominant (AD) Aortic valve disease 2 (MIM:614823) which features bicuspid aortic valve and aortic coarctation^51^. Three VUS cases of probable genetic etiology had death certificate phenotypes of congenital encephalopathy. The prevalence of moderate or severe encephalopathy is 16/10,000 newborns^52^. They were AD Developmental and epileptic encephalopathy (DEE) 11 (*SCN2A* c.877A>G, p.Thr293Ala, MIM:613721); DEE7 (*KCNQ2* c.46G>A, p.Gly16Arg, MIM:613720); and DEE27 (*GRIN2B* c.2749T>C, p.Ser917Pro, MIM:616139). DEE are associated with Sudden Unexpected Death in Epilepsy (SUDEP) and death as a complication of status epilepticus^10–17,20–22,25–28,45–47^.

In SUID cases with variants in genes associated with more than one disorder, the disorder with greatest SUID likelihood was selected. For example, heterozygous variants in titin (*TTN*) were associated with AD Dilated cardiomyopathy 1G (MIM: 604145, which features dysrhythmias and severe congestive heart failure in the first decade of life), and not *TTN*-associated Familial hypertrophic cardiomyopathy 9 (MIM:613765), Myofibrillar myopathy 9 with early respiratory failure (MIM:603689) or Tardive tibial muscular dystrophy (MIM:600334), which have later onset or are less severe^38^. WGS identified *TTN* genotypes in four SUID cases and was the only recurrent probable genetic etiology. Two SUID cases had probable genetic etiology findings in two loci each: One had AD *CHRNE*–Slow channel congenital myasthenic syndrome 4A (MIM:605809) and AD *SCN1A*–DEE6B (MIM:619317). The other had AD *ATP1A2*–DEE98 (MIM:619605) and AD *TCF12*–Craniosynostosis 3 (MIM:615314). In addition to the encephalopathies mentioned above, death certificate phenotypes narrowed the differential diagnosis in two cases: One had phenotypes R95 (SIDS) and congenital encephalopathy and WGS findings of *ATP1A2*-DEE98 (MIM: 619605)^10–16,20,25–30^; The other had phenotypes W75 (Accidental Suffocation and Strangulation in Bed) and abnormal chromosome morphology, and WGS identified Jacobsen syndrome (Chr 11q23.3-qter del; MIM: 147791). Approximately 20% of infants with Jacobsen syndrome die due to congenital heart disease, infections, or intracranial hemorrhage^53,54^.

Six (17%) of 36 SUID probable genetic etiologies had autosomal recessive MOI while the rest were AD. Thirty-three genotypes involved small variants and six were structural variants. These proportions were similar to prior infant cohorts with genetic diseases^12–17,25–29,33,34^. Thirteen (39%) small variants were absent from gnomAD v4.1 and 13 (39%) from ClinVar, which were were higher proportions than in other infant cohorts with genetic diseases, reflecting the novelty of WGS studies of SUID.

### Identification of additional G risks in individual SUID cases

Ninety-nine (47%) of 212 SUID cases had 149 WGS findings in 138 loci that were categorized as possible genetic etiology (Table S2). In contrast to SUID of probable genetic etiology (P/LP variants with *f* <8.2 × 10^−6^), these were 144 VUS genotypes with *f* <8.2 × 10^−6^ and 5 LP genotypes with *f* <3.6 × 10^−5^. The latter frequency was higher than supported by the prevalence of SUID and implied <25% penetrance. Twenty-nine SUID cases had >1 possible genetic etiology. Sixteen (12%) disorders were recessive, two were structural variants and 147 were nucleotide genotypes. Ninety-five (64%) variants were absent from ClinVar and 47 (32%) from gnomAD, which were also higher proportions than other infant cohorts with genetic diseases.

*In toto*, WGS identified a probable/possible genetic etiology in 121 (57%) SUID cases (Fig. 2a). Thirty-six had >1 disorder (G x G SUID risk), for a total genetic burden of 173 genetic risks (Fig. 2b). For example, one infant had four possible genetic disorders and one probable genetic disorder (*SMAD6*–Aortic valve disease 2, MIM:614823, probable genetic etiology; *BMP2*–Short stature, facial dysmorphism and skeletal anomalies with or without cardiac anomalies 1, MIM:617877; *GLI2*–Holoprosencephaly 9, MIM:610829; *NKX2-1*–Choreoathetosis, hypothyroidism and neonatal respiratory distress, MIM:610978; and *TRIO*–Intellectual developmental disorder with microcephaly 44, MIM:617061). Genetic heterogeneity was extreme: 144 unique loci were observed, of which 17 were observed twice, 4 three times, and one (*TTN*) 6 times. When disorders were categorized by principal organ affected, congenital anomaly syndromes were the most common (82 infants, 39%), followed by cardiac dysfunction (25, 12%) and seizure disorders (19, 9%; Fig. 2c).

**Fig. 2:**
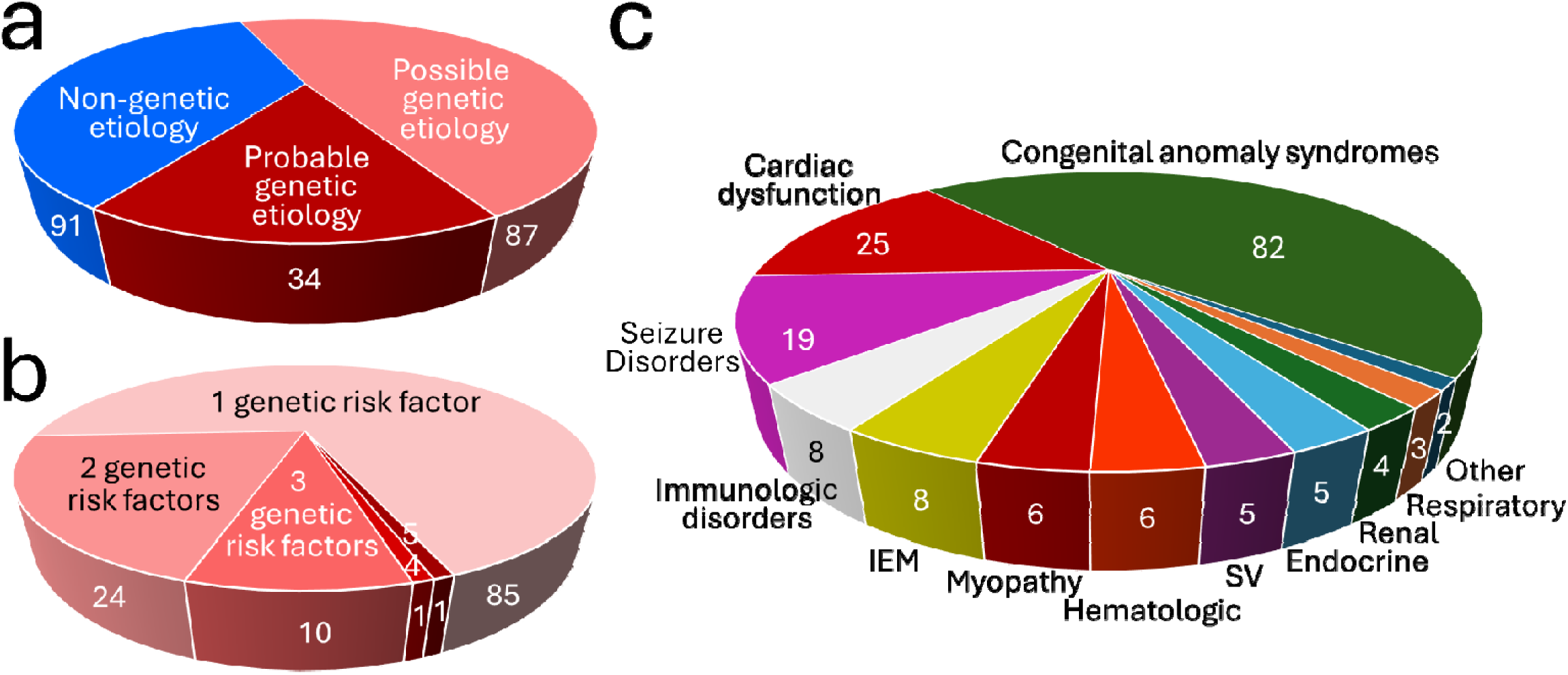
Pie charts showing categorization of 212 SUID cases by genetic etiology. **a.** Categorization of SUID etiology as possibly, probably, or not genetic based on WGS findings. **b.** Number of genetic risk factors identified by WGS in SUID cases with probable or possible genetic etiology. **c.** Categorization of SUID genetic risk by principal organ affected.

### Available therapeutic interventions for SUID cases with G risks

An important reason to identify SUID of genetic etiology is prevention by specific therapeutic interventions^16,17,33,34^. The 144 genetic disorders identified in SUID cases were categorized by presenting clinical features, natural history, appropriate confirmatory tests and specialist consultants, and the efficacy, evidence of efficacy, indications, contraindications, and urgency of initiation of available therapeutic interventions^35,55,56^. One hundred and twenty (83%) disorders had therapies that could prevent SUID if affected cases were identified at birth (Table S3).

### Quantification of SUID risk attributable to WGS findings

Based on graphical models of SUID G and E risk factors for 107 genetic disorders (Supplementary Methods, Table S1, Fig. 1, 3), probable/possible genetic etiologies were converted to SUID prevalence risk ratios (PRR) for each disorder and case (where >1 G risk was identified). A PRR of 991 represented 100% risk. The median PRR for the 36 disorders of probable SUID genetic etiology was 14.0 (range 3.0-792). The median combined PRR for the 34 SUID cases of probable genetic etiology, with 54 G risks, was 38.8 (range 3.0-991). For all 121 SUID cases of probable or possible genetic etiology, the median combined PRR was 10.0 (range 1.3-991). The distribution of PRR values allowed categorization of SUID cases into three groups: 12% of cases (26) had very strong G risk (PRR >99), 34% (72) had strong G risk (PRR 3.7 – 99), and 54% (114) had weak/no G risk (PRR 0 – 3.3)(Fig. S1, 4a).

The graphical models also informed representation of individual composite G x E SUID risk (Supplementary Methods, Fig. 1, 3). Of 107 SUID G risk factors, 64% were causally associated with congenital anomalies, 56% with respiratory distress syndrome, and 33% with premature birth (Table S1). Therefore, SUID risks in infants with strong/very strong G risk were conditioned on these factors. For example, Jacobsen syndrome in infant 1235114 (prevalence 1/100,000 births; infant mortality rate 20%; SUID PRR 198) was sufficient alone to account for SUID (Fig. 3a)^53,54^. A more complicated example was infant 1253290 with LP genotypes for both *CHD7*-CHARGE syndrome (prevalence 8/100,000 births; infant mortality rate 20%) and *NODAL*-Heterotaxy 5 (prevalence 1/100,000 births; infant mortality rate 23%; SUID PRR 15.1; Fig. 3b). CHARGE and heterotaxy syndromes are associated with congenital anomalies and respiratory distress syndrome, while heterotaxy is also associated with prematurity. In these infants, the effects of E risk factors were subsumed by G risks. These examples typified SUID cases with very strong G risk (G PRR >99) in which integrated G x E models of SUID risk conditioned for all E risk factors (Fig. 3e, Supplementary Methods).

**Fig. 3:**
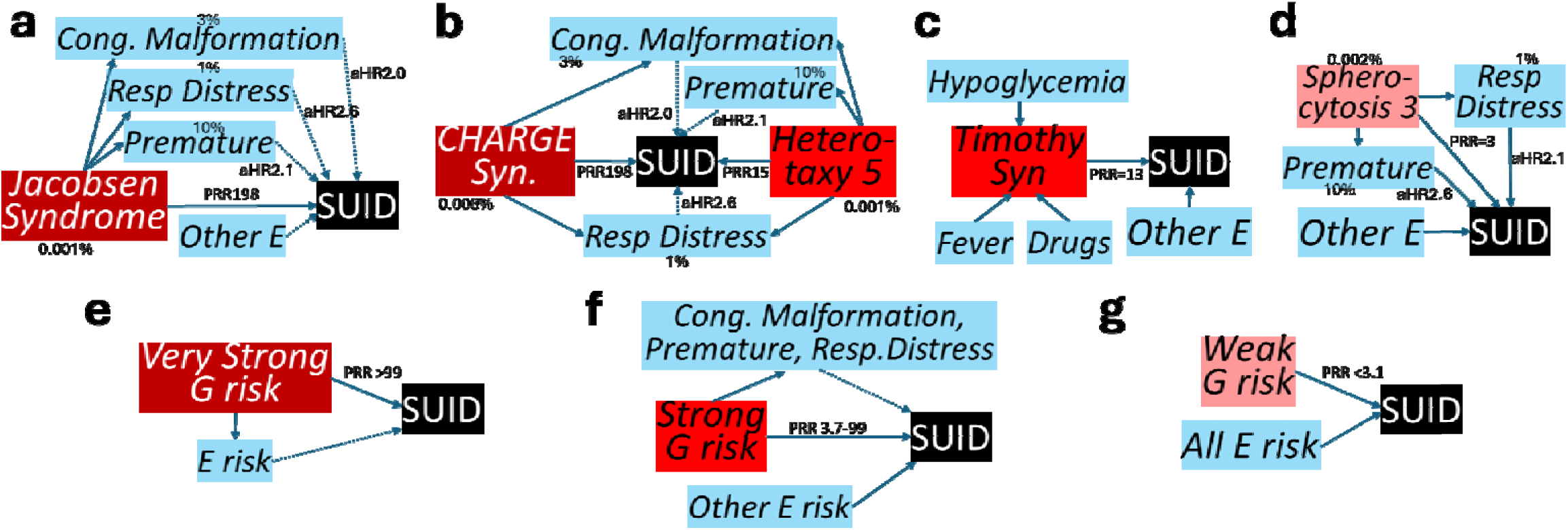
Graphs of integrated G x E models of causal association of SUID. Arrows with solid lines represent direct associations and those with dotted lines represent associations subsumed by a direct association. Birth prevalence (%) is indicated. Risk magnitudes are indicated by prevalence risk ratios (PRR) for G factors and cohort adjusted hazard ratios (aHR) for E factors (from Table 1). **a** Infant 1235114 with Jacobsen syndrome, a very strong SUID risk factor, considered sufficient for SUID alone. It is causally associated with congenital anomalies, respiratory distress syndrome, and premature birth. E SUID risk factors were considered subsumed to Jacobsen syndrome for SUID causality. **b** Infant 1253290 with two very strong SUID risk factors (CHARGE syndrome, Heterotaxy 5). Both are associated with congenital anomalies and respiratory distress syndrome, and Heterotaxy with premature birth. These and all other E SUID risk factors were considered subsumed to the G factors for SUID causality. **c** Infant 1229840 with a strong SUID risk factor (a VUS in *CACNA1C*, the causal gene in Timothy syndrome). The E factors fever, infection, hypoglycemia and various drugs, modify Timothy syndrome-associated SUID risk. **d** Infant 1253265 with Spherocytosis 3, a weak SUID risk factor that is associated with premature birth and respiratory distress. However, given the relatively weak G risk, all E factors were retained. **e-g** Causal G x E models for SUID cases with very strong (**e**), strong (**f**), and weak (**g**) G risk.

In contrast, in the 34% SUID cases with strong G risk (PRR 3.7 – 99), E factors frequently were potential modifiers of direct G risks, and G x E interactions led to SUID. For example, infant 1229840 had a VUS in the α1C subunit of the L-type, voltage gated Ca^2+^ channel (*CACNA1C,* Fig. 3c). Pathogenic variants in *CACNA1C* cause Timothy syndrome (TS, MIM:601005) or Long QT syndrome, Supplementary Methods). SUID occurs in 15% of TS infants via dysrhythmias and sudden cardiac death (PRR 159). However, the PRR attributable to the VUS in this infant was 13 (square root of PRR for P variants). E factors such as fever, infection, hypoglycemia and various drugs alter Ca^2+^ channel dynamics to trigger dysrhythmias in TS. These E factors were considered intermediates in a causal TS – SUID chain that acted to modify G risk. TS exemplified SUID cases with strong G risk in which an integrated G x E model of SUID risk conditioned for selected E risk factors (Fig. 3f, Supplementary Methods).

The remaining 54% SUID cases had weak/no G risk (PRR 1 – 3.3). SUID E risk factors predominated in these cases. For example, infant 1253265 had a P genotype for *SPTA1*-Spherocytosis 3 (prevalence 2/100,000, Fig. 3d). Infant death, due to severe anemia, is very uncommon (SUID PRR 3). While Spherocytosis is associated with premature birth and respiratory distress, the relative effect sizes of these E risks were of similar magnitude to that conferred by G risk. This case exemplified SUID cases with weak G risk in which an integrated G x E model of SUID risk retained all E factors (Fig. 3g).

### Analysis of maternal, pregnancy, or infant risk factors for SUID with and without cases with strong G risk

We compared maternal, pregnancy and infant characteristics of the 212 SUID cases and 620,392 infants alive at age 1 year from 2005-2018 to identify SUID E risk factors (Fig. S1). Thirteen of 24 maternal factors differed significantly between groups (Table 1). SUID risk was significantly lower in mothers of Hispanic ethnicity, those aged ≥34 years, and those with pre-existing diabetes mellitus. SUID risk was significantly higher in mothers of Black race, those with educational attainment <12^th^ grade, those who were overweight or with obese body mass index, inadequate prenatal care, Medicaid as healthcare payor, depression, nicotine, cannabis related diagnosis, other substance related diagnosis, and pre-existing hypertension. Four of five pregnancy or infant characteristics differed significantly between groups (Table 1). SUID risk was higher in multiparous gestation, preterm delivery, very preterm delivery, and respiratory distress syndrome.

In comparison to an identical analyses of 211 SUID cases and 484,698 infants alive at age 1 year from 2005-2017^18^, the larger cohort herein enabled four factors to reach significance (Hispanic ethnicity, Black race, age ≥35 years and depression). The largest change in risk between the 2005-2017 and 2005-2018 datasets was in cannabis related diagnosis, which increased from aHR 2.7 (CI 1.5,5.0) to 3.7 (CI 2.1,6.7). These cohorts differed both by inclusion of the year 2018 and by removal of SUID cases without archived dried blood spots.

Informed by knowledge of SUID G risk, we re-evaluated E risks (Fig. S1). First, we removed the 26 SUID cases with very strong G risk and compared the remaining 186 SUID cases with 620,392 infants alive at 1 year of age. We hypothesized that removal of cases with very strong G risk, in which whom E risk factors were not considered contributory to SUID, should decrease effects of E factors explained by G risk and increase effects of E factors unrelated to G risk. Of 16 significant E factors in comparison with all SUID cases, one factor (respiratory distress syndrome) lost significance (Table 1). In addition, substantial changes in magnitude were observed for three factors: Risks attributable to cannabis related diagnosis increased from aHR 3.7 to 4.4, other substance related diagnosis increased from aHR 2.6 to 3.0, and very preterm delivery decreased from aHR 3.4 to 2.8.

Next, we removed the 98 SUID cases with strong/very strong G risk and evaluated E risk factors in the remaining 114 SUID cases with weak/no G risk (Fig. S1, 4b). We hypothesized that removal of these cases should further decrease effects of E factors explained by G risk and increase effects of E factors unrelated to G risk. Of 16 significant E risks identified in comparison with all SUID cases, six lost significance: Obesity, inadequate prenatal care, public payer, respiratory distress syndrome, very preterm delivery, and preexisting hypertension (Table 1). In addition, the magnitude of risk changed appreciably for three factors: Risk attributable to black race increased from aHR 1.9 to 2.5, cannabis related diagnosis increased from aHR 3.7 to 6.0, and for other substance related diagnosis increased from aHR 2.6 to 3.5 (Fig. 4b).

**Figure 4:**
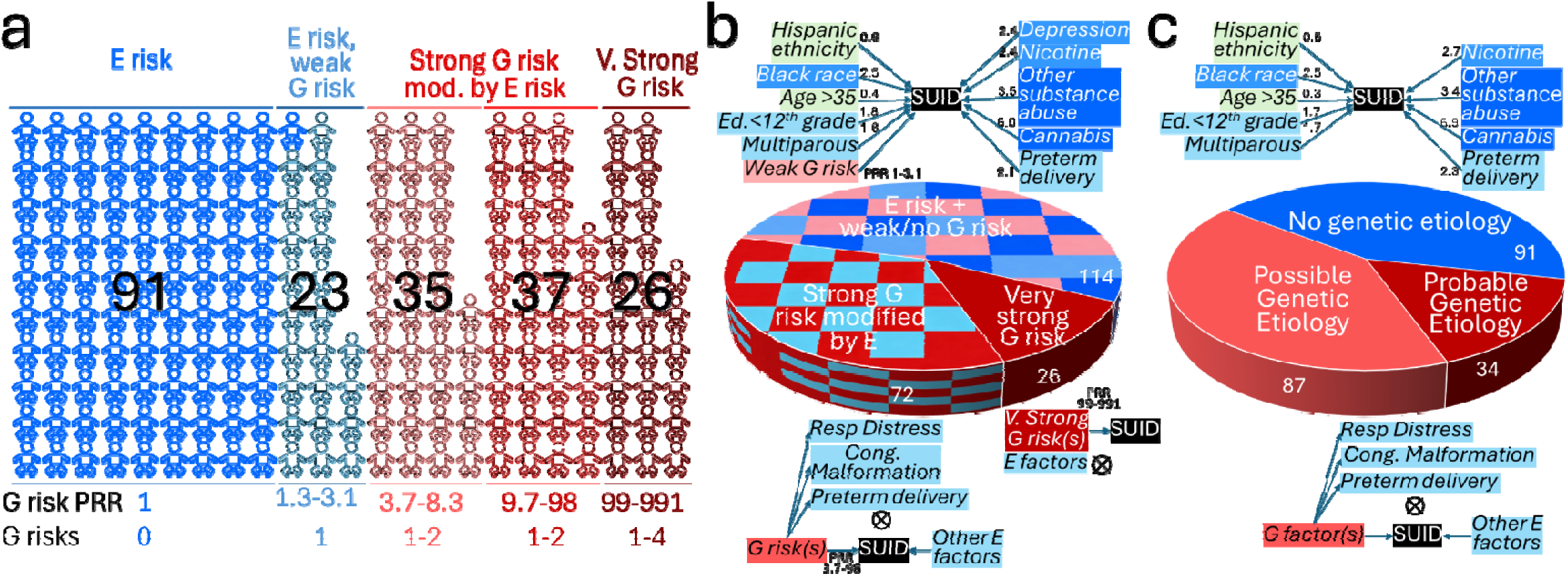
Categorization of 212 SUID cases by the magnitude of genetic (G) and environmental (E) risks. **a** Categorization by G risk Prevalence Risk Ratio (PRR). **b** Pie chart showing integrated G*E models for SUID cases with weak/no G risk, strong G risk, and very strong G risk. The magnitude of individual E risks is shown as adjusted hazard ratios. **c.** Pie chart showing integrated G*E models for SUID cases without genetic etiology, possible genetic etiology and probable genetic etiology.

In a sensitivity analysis, we re-evaluated E risk factor effects following removal of the 125 SUID cases with probable and possible genetic etiology (Fig. S1, 4c). Of 16 significant E risks identified in comparison with all SUID cases, five lost significance: Inadequate prenatal care, public payer, respiratory distress syndrome, very preterm delivery, and preexisting hypertension (Table 2). Likewise, the magnitude of risk increased substantially for black race (aHR 1.9 to 2.5), cannabis related diagnosis (aHR 3.7 to 6.9) and other substance related diagnosis (aHR 2.6 to 3.4; Fig. 4c).

**Table 2:**
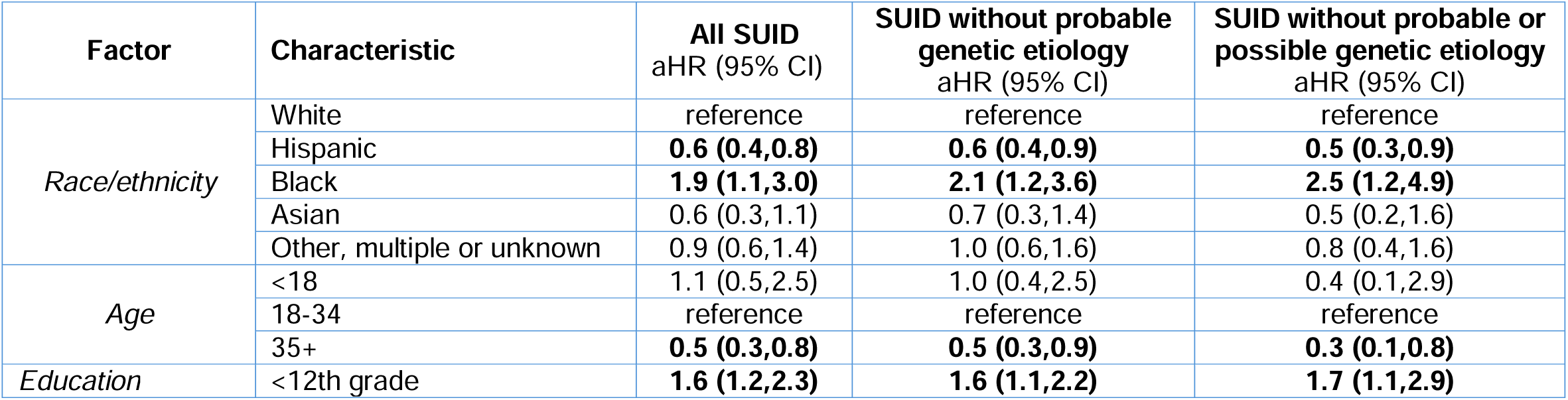
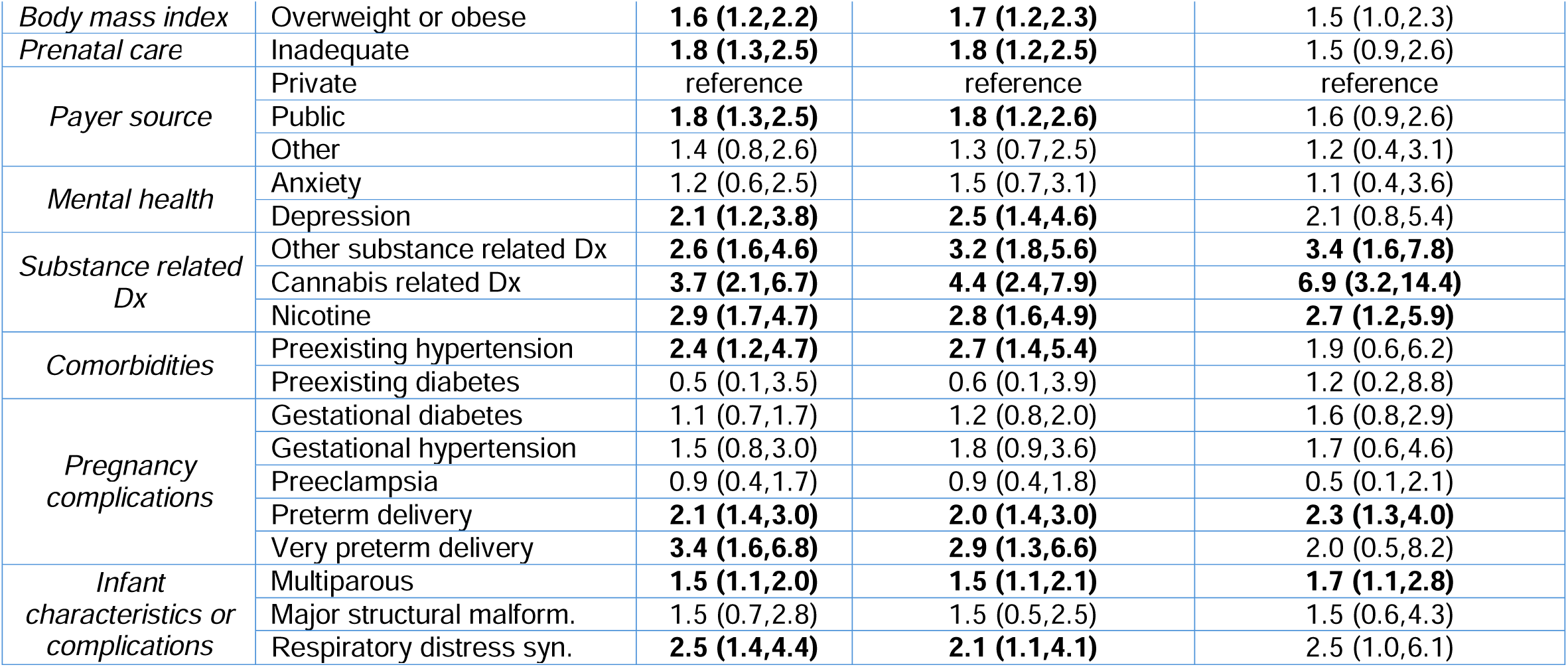
Adjusted hazard ratios (aHR) and 95% confidence intervals (CI) for maternal, infant and environmental characteristics of SUID cases compared to 620,392 infants alive at age 1 year. Comparison groups were all SUID cases (n=212), SUID cases without probable genetic etiology (n=178), and SUID cases without probable or possible genetic etiology (n=91). Models adjusted for maternal race/ethnicity, age, education, body mass index, prenatal care, payer source, and smoking. Each factor from individual models. aHR in bold were significant (p<.05).

## Discussion

The premise of this study was that multifactorial G x E analysis of SUID risk would overcome limitations of single domain epidemiology studies, unveil or recalibrate G and E risk factor effects, and suggest preventative interventions that have hitherto not been emphasized (Fig. S1). This premise was confirmed, yielding three key insights. Firstly, the etiology of SUID was heterogeneous and three groups were evident, 12% with very strong G risks, 34% with strong G risks modified by E risks, and 54% with E risks and weak or no G risk (Fig. 4b). Secondly, the causal relevance of E risks differed in these groups. Finally, the analysis suggested two preventative interventions – newborn screening by WGS to identify and ameliorate G risk and emphasis of cannabis avoidance in educational campaigns for SUID. These conclusions were supported by two different analysis strategies.

WGS identified G risks in 57% of SUID cases. This proportion agreed with a single similar study, which identified G risks in 65% of 144 SUID cases by WGS.^25^ It was also consistent with other genomic sequencing studies in SUID or other infant deaths^11–17,21,22,25–28^. The occurrence of G risk in over one half of SUID cases implies that preventative strategies that ignore such risks will, at best, be only partially effective.

G risks were extremely heterogeneous, with 144 loci in 121 SUID cases, of which 85% were unique to a single patient. This finding concurred with prior WGS studies of SUID and infant death in non-consanguineous populations.^16,17,25^ Therapies with potential to prevent SUID were identified for 83% of G risk loci. An implication of extreme G risk heterogeneity is that preventative strategies will require comprehensive identification methods, such as newborn screening by WGS, allied with population precision health delivery^35,55,56^. Research studies worldwide are exploring use of WGS-based newborn screening for hundreds of severe genetic diseases with effective treatments. The loci identified herein showed considerable overlap with those identified in a single similar study (25% locus overlap encompassing 38% of SUID cases herein).^25^ Recurrent loci were *TTN*-dilated cardiomyopathy 1G (5.3% of 356 total SUID cases), *TSC1*-tuberous sclerosis (1.7% of all cases), *RYR2*-catecholaminergic polymorphic ventricular tachycardia (2.0%), *DSP*-dilated cardiomyopathy (1.7% total cases), *NOTCH1*-Adams-Oliver syndrome 5 (1.4%), *CACNA1H*-familial hyperaldosteronism IV (1.4%), *ANK2*-Long QT syndrome 4 (1.4%), and *FLNA*-Melnick-Needles syndrome (1.1%)^25^. The overlap in G risk increased when extended to gene families. For example, 3.7% of 356 total SUID cases had G risks in voltage-gated sodium channels. Previously we showed that genetic disease loci differed between infant deaths and severity-matched infant survivors^16^. In that cohort, the specific genetic etiology had prognostic value (positive predictive value for death 85%)^17^. Thus WGS may be effective for newborn screening for SUID risk.

Despite G risk heterogeneity, several previously reported unifying SUID mechanisms were evident.^10–17,20,21,25–30^ One third of G risks featured seizures and conferred risk of SUDEP or status epilepticus, 39% were cardiac disorders associated with arrhythmias, sudden death or acute heart failure. Other prominent etiologies were immunodeficiencies (12%) and breathing or respiratory disorders (22%). These groups represent relatively homogeneous SUID mechanisms which converge on potential preventative or therapeutic interventions^10–16,21,22,25–28^.

G risks were categorized in two ways – by likelihood of being affected by a genetic disease known to cause infant death and by magnitude of SUID risk. Likelihood of disease expression was classified as either SUID of probable genetic etiology (high likelihood of expression of a disease associated with infant mortality) or possible genetic etiology (lower likelihood of expression). Sixteen percent of SUID cases were of probable genetic etiology, defined as P/LP genotypes in genes known to cause infant death and with population frequencies consistent with the low prevalence of SUID (Fig. 3a). This rate was similar to a non-overlapping study of WGS in 112 infant deaths in a hospital system in San Diego County (25% diagnoses)^17^. Classification as probable genetic etiology was not quite the same as molecular diagnoses, since it excluded 5 diagnostic findings that exceeded the frequency cutoff (Klinefelter syndrome and four cases demoted to possible diagnoses, Supplementary Discussion). Including these five cases, the solved rate herein (19%) was similar to WGS studies with broad NICU enrollment^59,60^.

In diagnostic WGS interpretation, cases with variants of uncertain significance (VUS) are considered unsolved. VUS are not benign, however, and confer risk of disease albeit of uncertain magnitude. Herein, VUS findings were considered possible genetic etiologies. They were defined as VUS genotypes in genes known to cause infant death and with population frequencies consistent with SUID prevalence. We included the four cases mentioned above that had P/LP genotypes and a slightly higher frequency. A possible genetic etiology was identified in 48% of SUID cases. There was more than one WGS finding in 30% of SUID cases, which was higher than reported for diagnostic WGS in individuals with suspected genetic diseases (Figure 2b)^61^. Many variants identified were novel, which often betokens *de novo* occurrence, and was consistent with the purifying hyperselection conferred by SUID^35^.

Literature review indicated that the risk of infant death conferred by genetic disorders varied 500-fold (Supplementary Methods). In order to perform integrated G x E SUID risk analysis we converted WGS findings from qualitative probability of being affected by a genetic disorder to quantitative SUID risk on a scale that allowed comparison with E risk. Significant E risk hazard ratios herein varied from 1.5 to 6.9. We utilized Prevalence Risk Ratios (PRR) to convert genetic disease likelihood to individual SUID G risk category. PRRs were calculated for each genetic disorder, each expressivity likelihood (probable or possible genetic etiology), and each SUID case (where there was more than one WGS finding). The PRR was the literature value for infant mortality rate for each genetic disorder divided by the population prevalence of SUID (101/100,000)^8^. While PRR values were approximate and had caveats (Supplementary Discussion), they informed categorization of G risk for each SUID case as very strong (combined PRR >100), strong (3.7-99), or weak (<3.7). These categories were reasonably robust: While 12.3% of SUID cases had strong G risk, decreasing the cutoff from PRR >100 to >50 increased it only to 15.6%. Likewise, the proportion of SUID cases with weak risk (53.3%) was unchanged between PRR cutoffs of 3.2-3.7.

Categorization of G risk informed integrated understanding of SUID risk. In the 12.3% of SUID cases with very strong G risk, E risk was relatively unimportant for SUID outcome (Fig. 3c). Preventative strategies for such cases should include identification and amelioration of specific genetic disorders. In the 34.4% of SUID cases with strong G risk, it alone was generally insufficient for SUID outcome. Instead, E risk factors were considered causal modifiers of G risk. For example, the death certificate of infant 1261156 listed congenital encephalopathy. This infant had voltage-gated potassium channel *KCNQ2*-Developmental and epileptic encephalopathy 7 (MIM:613700, PRR 14), a probable genetic etiology. Neuronal K^+^ channel gating is modified by a variety of E factors, such as fever, hypoxia, hypoglycemia, anxiety, and various drugs^62–66^. DEE7-associated SUID is preventable by early diagnosis and seizure control with targeted anti-seizure medicines. Most genetic syndromes identified by WGS included congenital anomalies, preterm birth, and respiratory distress as features. Depending on the genetic disorder, the E factors preterm delivery (aHR 2.1), very preterm delivery (aHR 3.4), major structural malformation (aHR not significant), and respiratory distress syndrome (aHR 2.5) were either confounders or part of the causal pathway. Preventative strategies for strong G risks, should include identification and amelioration of specific genetic disorders together with minimization of E risks.

We previously reported results of multi-domain SUID E risk in all San Diego infants. Herein we expanded that analysis by one year^18^. The addition of 2018 data modestly increased E factor effect sizes to the extent that five factors reached significance (maternal Hispanic ethnicity decreased from aHR 0.8 to 0.6, maternal Black race increased from aHR 1.6 to 1.9, maternal age >35 years decreased from aHR 0.7 to 0.5, and maternal depression increased from 1.8 to 2.1). The largest change in E factor effect size, however, was in prenatal cannabis related diagnosis, which increased from aHR 2.7 to 3.7. It should be noted that while recreational cannabis use was legalized in California at the end of 2016, commercial sales began in 2018. Thus, broad cannabis availability was associated with increased SUID risk. Furthermore, the aHR associated with cannabis further increased (from 3.7 to 4.4) upon removal of cases with very strong G risk (in whom E risks were relatively unimportant). Upon removal strong and very strong G risk SUID cases, the cannabis aHR increased to 6.0. This was recapitulated in a sensitivity analysis, when removal of cases of probable and possible genetic etiology increased cannabis aHR to 6.9. Maternal prenatal cannabis use was the strongest risk factor for SUID. Identical trends were observed for other maternal substance abuse upon conditioning for genetic risks. Likewise, upon removal of strong and very strong G risk, SUID risk attributable to black race increased from aHR 1.9 to 2.5. This was also recapitulated in a sensitivity analysis. Thus, increased SUID risk in black infants was not explained by rare genetic variants. As expected, upon conditioning for cases explained by strong or very strong G risk, E risks of factors explained by G risk (respiratory distress syndrome and very preterm birth) became insignificant. Finally, in 53.3% of SUID cases G risks were weak or absent. In such cases preventative strategies should focus on amelioration of the eight remaining significant E risks.

This study had limitations. While highly comprehensive, short-read WGS will have underestimated G risk associated with structural variations, particularly insertions, balanced translocations or complex variations, and genes that are challenging to sequence, such as those with highly homologous pseudogenes or repeat expansions^33,34,57,58^. WGS was limited to SUID cases, which prevented direct case-control comparisons for WGS. The methods used for conversion of WGS findings to quantitative risk had not previously been validated. Only 212 SUID cases were examined, which is few relative to the etiologic heterogeneity of SUID. We used administrative data (ICD codes and birth records) for demographic factors and comorbidities which can lead to misclassification. However, such misclassification should be non-differential and bias results towards the null hypothesis. Substance-related diagnoses are known to be under-reported in hospital discharge summaries and likely represent disordered use. We only captured infants who died in California in the first year of life. Infants who left the state were assumed alive at one year, which may not have been the case. We were unable to adjust standard errors for non-independence of siblings. Death certificates are known to be incomplete and inaccurate. We did not have information on sleep position, a well-established risk factor for SUID. Nevertheless, inclusion of almost the entire population of infants in San Diego County over a 13-year period, an ethnically and economically diverse sample, increases the likelihood that results are generalizable to other similar populations.

In summary integrated G x E analysis expanded understanding of risk factors for SUID. Genetic risk was identified in 57% of SUID cases and was associated with median PRR of 10. While genetic risks were highly heterogeneous, they were readily identified by WGS, and a set of recurrent loci was apparent. Of note, 83% of genetic risks had available preventative or therapeutic interventions, suggesting that newborn diagnosis and precision intervention could lead to substantial reduction in SUID rates. For example, population newborn screening for these disorders by WGS could substantially reduce SUID rates. Upon conditioning for G risks, the magnitude of several E risks increased. Prenatal maternal cannabis was the largest E risk factor. Of note, the broad availability of cannabis in 2018 was associated with increased SUID risk. Educational campaigns for SUID should emphasize perinatal cannabis avoidance.

## Declaration of interests

Erwin Frise is an employee and shareholder of GeneDx Inc. Mark Yandell is a shareholder of GeneDx Inc.

## Acknowledgments

This manuscript was supported by NIH grant grant R01HD101540. The California Department of Public Health is not responsible for the results or conclusions drawn by the authors of this publication. *A Deo lumen, ab amicis auxilium*.

## Web resources

Genome to Treatment (GTRx) is available at https://gtrx.rbsapp.net/. Information about the BeginNGS consortium is available at https://radygenomics.org/begin-ngs-newborn-sequencing/.

## Data and code availability

There are restrictions to the availability of raw individual data due to data privacy and confidentiality laws. Anonymized and pseudonymized individual data generated in this study, subject to the terms of informed written consent documents, and state and federal laws, are provided in the Supplemental Information.

GTRx and the GTRx REDCap instance is available at https://gtrx.rbsapp.net/ and code is available from Christian Hansen and at https://github.com/rao-madhavrao-rcigm/gtrx. The DRAGEN Platform and Illumina Connected Analytics are available from Illumina (Shyamal Mehtalia). GEM and Transformer are available from Fabric Genomics.

## Notes

### Competing Interest Statement

Erwin Frise is an employee and shareholder of GeneDx Inc. Mark Yandell is a shareholder and consultant of GeneDx Inc.

### Author Declarations

The study was approved by the Committee for the Protection of Human Subjects within the California Health and Human Services Agency and the University of California San Diego Institutional Review Board.

